# Evidence of widespread endemic populations of highly multidrug-resistant *Klebsiella pneumoniae* seen concurrently through the lens of two hospital intensive care units in Vietnam

**DOI:** 10.1101/2021.06.29.21259521

**Authors:** My H. Pham, Le Thi Hoi, Mathew A. Beale, Fahad Khokhar, Nguyen Thi Hoa, Patrick Musicha, Grace Blackwell, Hoang Bao Long, Dang Thi Huong, Nguyen Gia Binh, Dao Xuan Co, Tran Giang, Cuong Bui, Hai Ninh Tran, James Bryan, Archie Herrick, Theresa Feltwell, Behzad Nadjm, Julian Parkhill, H. Rogier van Doorn, Nguyen Vu Trung, Nguyen Van Kinh, M. Estée Török, Nicholas R. Thomson

## Abstract

**Background:** Extended spectrum beta-lactamases-producing (ESBL-P) and/or carbapenem-resistant (CR) *Klebsiella pneumoniae* have severely restricted available treatment options in healthcare settings in Vietnam. Understanding the diversity and transmission mechanisms of ESBL- and carbapenemase-encoding *K. pneumoniae* is important in both hospital and community settings for patient management.

**Methods:** We conducted a 6-month prospective cohort study of 69 Intensive care unit (ICU) patients from two hospitals in Hanoi, Vietnam. Longitudinally collected samples from patients and the ICU environment were cultured on selective media, and 357 *K. pneumoniae* colonies were whole genome sequenced. We performed phylogenetic analyses, and correlated phenotypic antimicrobial susceptibility testing with genotypic features of *K. pneumoniae* isolates. We constructed transmission networks of patient samples, relating ICU admission times and locations with genetic similarity of infecting *K. pneumoniae*.

**Findings:** Despite being geographically and clinically separated, the two hospitals shared closely related strains carrying the same array of antimicrobial resistance genes. Many patients carried the same resistant *K. pneumoniae* clone from admission to discharge. 45.9% of total isolates carried both ESBL- and carbapenemase-encoding genes, with high minimum inhibitory concentrations. We found a novel co-occurrence of *bla*_*KPC-2*_ and *bla*_*NDM-1*_ in 46. 6% of samples from the globally successful ST15 lineage.

**Interpretation:** These results highlight the high prevalence of ESBL-positive carbapenem-resistant *K. pneumoniae* in Vietnamese ICUs. Through studying *K. pneumoniae* ST15 in detail, we illustrated how important resistance genes are coalescing in stains carried broadly by patients entering the two hospitals directly or through referral.

**Funding:** This study was supported by the Medical Research Council Newton Fund, United Kingdom (grant MR/N029399/1); the Ministry of Science and Technology, Vietnam (grant HNQT/SPÐP/04.16); This research was funded in whole by the Wellcome Trust (grant 206194). For the purpose of Open Access, the author has applied a CC BY public copyright licence to any Author Accepted Manuscript version arising from this submission.

## INTRODUCTION

In healthcare settings, *Klebsiella pneumoniae* causes approximately one-third of all Gram-negative bacterial infections [1], ranging from pneumonia and urinary tract infections to sepsis, with a mortality rate as high as 70% [2]. Furthermore, many *K. pneumoniae* lineages are multiply resistant to first-line and last-line drugs, including aminoglycosides, fluoroquinolones, cephalosporins and carbapenems, via the production of extended-spectrum beta-lactamases (ESBLs). The ability of *K. pneumoniae* to become multidrug-resistant (MDR) is linked to its global clinical success and dissemination. Indeed, five clonal groups are responsible for 72% of all reported outbreaks globally [3]. Consequently, MDR *K. pneumoniae* have severely limited treatment options in most clinical settings, even for routine infections, but especially for patients in intensive care units (ICUs) [1, 3, 4]. As antimicrobials are the primary treatment, resistance is also associated with increased treatment costs, prolonged durations of infection, more complex sequelae, and elevated mortality rates [5].

After China, Vietnam has the second-highest prevalence of ESBL producing *K. pneumoniae* globally, accounting for 67% of all isolates causing urinary tract infections (UTIs) [6]. The proportion of carbapenem resistance was reported to be between 1 and 17% in South and South-East Asia, and less than 10% in Vietnam in 2010 [7, 8]. Moreover, the prevalence of carbapenem-resistant *K. pneumoniae* has seen recent and sustained increases in Vietnam, reaching 24% in 2016 [9].

Here, we focussed on ESBL-producing and/or carbapenem-resistant *K. pneumoniae* isolates to understand how they, and the drug resistance genes they carry, were circulating in Vietnamese hospital settings. We sampled patients admitted to the ICUs of two geographically close but independent hospitals in Hanoi: The National Hospital for Tropical Diseases (NHTD) a leading hospital for infectious and tropical diseases in northern Vietnam, and Bach Mai Hospital (BMH) a multi-speciality and tertiary referral hospital. Whilst these hospitals are adjacent to each other, each hospital has independent facilities and staff, and patients are not usually transferred between them.

## METHODS

### Study setting and participants

The NHTD ICU has 22 beds and receives up to 400 admissions per year. The BMH ICU has 45 beds with up to 1,200 admissions per year. All patients aged over 18 years old who were admitted between 1^st^ June 2017 and 31^st^ January 2018 were eligible for inclusion in a prospective surveillance study. Patients who were admitted for longer than the mean length of stay (20 days in NHTD ICU and 10 days in BMH ICU) and grew *Klebsiella pneumoniae* from their clinical samples were enrolled in this study.

### Study procedures

Written informed consent was obtained from the patient or their relative. Clinical data were collected into a case record form and entered into the study database. Clinical samples (stool or faecal swabs, sputum or tracheal aspirates, urine, and wound swabs) were collected from each patient on admission to ICU, at weekly intervals, and on discharge from ICU. Environmental samples were collected monthly using flocked swabs from individual patient bed spaces (including bed rails, tables, ventilation equipment, door handles, and taps) and pooled before plating on selective media.

### Laboratory Methods

Clinical and environmental specimens were cultured on CHROMagar™ ESBL or CHROMagar™ mSuperCARBA™ selective media (CHROMagar, France). Single colony picks were identified using matrix-assisted laser desorption-ionisation time-of-flight mass spectroscopy machine (MALDI-TOF MS, Brucker Diagnostics, Germany). Phenotypic antimicrobial resistance profiles were determined using VITEK®2 AST N350 card (Biomérieux, Marcy L’Étoile, France) and reported as resistant, intermediate or susceptible based on the Advanced Expert System [10]. Whole genome sequencing (WGS), Quality Control (QC) analysis, phylogenetic inference, comparative genomics and statistical analysis was performed as describe in detail in Supplementary Methods.

## RESULTS

### The relationship between *K. pneumoniae* isolates and AMR profiles seen in BMH and NHTD

Clinical specimens were collected from 69 patients admitted to NHTD (n=32) and BMH (n=37) ICUs and we also included 34 environmental samples collected from patients’ bed spaces (NHTD n=21; BMH n=13). In total, 357 *K. pneumoniae sensu stricto* isolates were cultured on selective media and successfully sequenced (Supplementary Data Table S1). Of the total ICU patients, 90.62% (n= 29) and 75.67% (n= 28) were referred to NHTD and BMH, respectively, from other hospitals. NHTD received nine patients from central hospitals, 19 patients from provincial hospitals and two from district hospitals. Most patients (n= 18) in BMH came from central hospitals, followed by nine from provincial and one from district hospitals. On the other hand, two and nine patients were admitted directly to NHTD and BMH, respectively. Patients are rarely transferred between NHTD and BMH and none of the patients included in this study had been transferred.

The WGS data were used to infer the phylogeny of all isolates using a core SNP alignment of 10,849 variant sites (Supplementary Figure 1) and to predict the sequence type of all isolates using *in silico* Multi locus Sequence Typing (MLST). Our collection comprised of 22 STs of which 88.23% (n=315) of isolates belonged to only five major clonal lineages: ST15 (36.69%, n=131), ST16 (18.76%, n=67), ST11 (14.00%, n=50), ST656 (12.32%, n=44) and ST147 (6.44%, n=23) (Supplementary Table 1).

There were 95 different phenotypic AMR profiles amongst the *K. pneumoniae* isolates with all isolates being resistant to at least one beta-lactam antibiotic, consistent with the use of ESBL selective media (Supplementary Data Sheet 2). 170 ESBL-positive isolates were also resistant to carbapenems (86.47% to ertapenem and 85.23% to meropenem; Supplementary Figure 2). These phenotypic resistance profiles were mostly concordant with those inferred from the WGS (Supplementary Figure 3): the largest disagreements were seen for isolates carrying either *bla*_SHV-12_ or *bla*_*OXA-48*_ where 16.70% or 26.7% of isolates were susceptible to cefoxitin or meropenem, respectively. Five isolates were phenotypically resistant but lacked all known ESBL (n=4) or CR (n=1) genes (Supplementary Data Sheet 1).

Isolates carrying *bla*_CTX-M-like_ genes were the most common ESBL-producers, accounting for 54.06% (n=193) of observed resistance, with *bla*_CTX-M-15_ (n=120) being the most common variant. Similarly, *bla*_KPC-2_ explained 51.54% (n=184) of the carbapenem resistance among the isolates (Supplementary Table 2). Overall, 63.86% (n=228) of *K. pneumoniae* isolates carried at least two, and up to a maximum of four different ESBL- and carbapenemase-encoding genes, with 45.93% of isolates (n=164) carrying genes encoding both.

Combining the above, multinomial-logistic regression analysis was used to determine if there was an association between AMR profile and/or ST and hospital (Figure 1). We observed a weak negative association between ST147 and NHTD (log odds ratio (OR)= −0.79, 95% confidence interval CI= [-1.72, 0.096]), but no association between any of the other STs and hospital of origin (Figure 1A). Equally, neither the range and richness of the 95 different phenotypic resistance profiles (Figure 1B) nor the distribution of each unique ST-ESBL-R/CR gene profile combination (plotted for the five major *K. pneumoniae* STs, Supplementary Figure 4) showed a strong association with hospital of origin, with no apparent effect from the length of hospital admission. Noted were minor differences in the pattern of AMR gene carriage within the same ST isolated from the two hospitals (Supplementary Figure 4).

**Figure 1:**
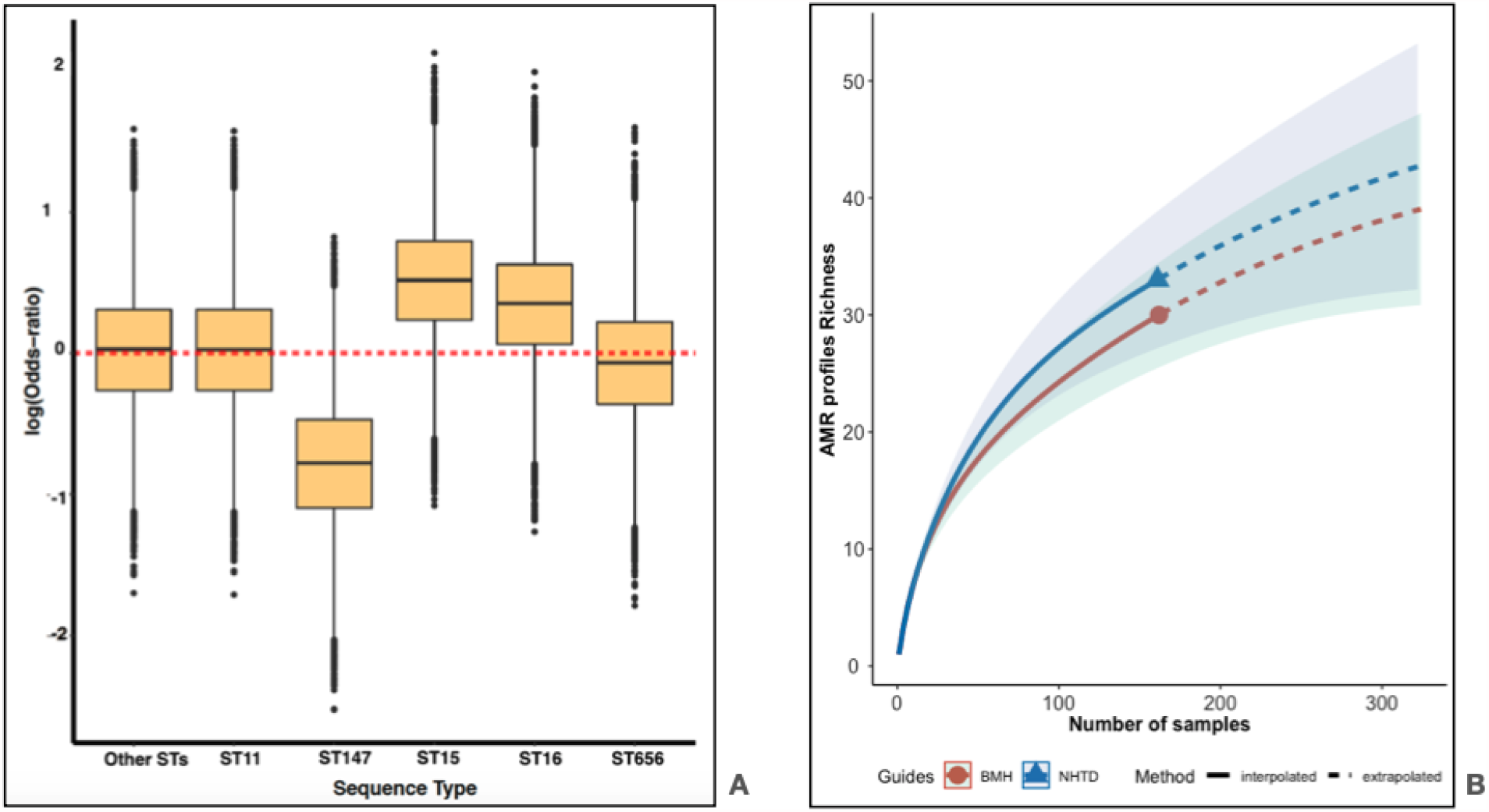
Association between ST and hospital, and the diversity of phenotypic AMR profile BMH and the NHTD. (A): Posterior density of odds ratio (on a log scale), from a multinomial regression of ST (dependent variable) and hospital (independent variable; reference group=BMH) after adjusting for within-patient sampling of STs with same antimicrobial resistance profiles. The horizontal axis shows five dominant STs and a group of other unrelated STs that were identified sporadically. The vertical axis indicates the log (Odds-ratio). In which, the closer the values of each whisker’s boxes to 0, the weaker association between STs and either one of the two hospitals. (B): Rarefaction curves, with 95% confidence intervals (95% CIs), indicated that the range and richness of resistance profiles is similar between two hospitals. The 95% CI*s* are shown o the graphs as the darker shaded areas behind the curve lines, green for BMH and blue for NHTD.

### Phylogenetic relationships of *K. pneumoniae* taken from individual patients

Considering the genetic uniformity of isolates spread across both hospitals, we sought to determine the patterns of *K. pneumoniae* carriage and transmission using the sequenced ST15 (the most numerous ST) genomes as an exemplar. We had 119 *K. pneumoniae* ST15 isolates cultured from 30 patients (16 from NHTD and 14 from BMH), and 12 corresponding environmental isolates (Figure 2). rPinecone was used to subdivide ST15 into 21 subtypes ([11]; ST15-1 to ST15-21) which combined with a ≤25 SNPs threshold range to infer transmission [7, 12], was used to plot a pairwise patient linkage network (Figure 2). This revealed three distinct patterns of transmission or carriage. Firstly, eight patients (marked as 1 to 8 in Figure 2; Pattern 1 – ‘persistent carriage’), four from each hospital, carried a single ST15 subtype throughout their admission. Here, a median of 7 SNPs (range 1 to 20 SNPs) differentiated sequenced isolates from the same patient, and a median of 43 SNPs differentiated isolates from their closest branching neighbour taken from different patients.

**Figure 2:**
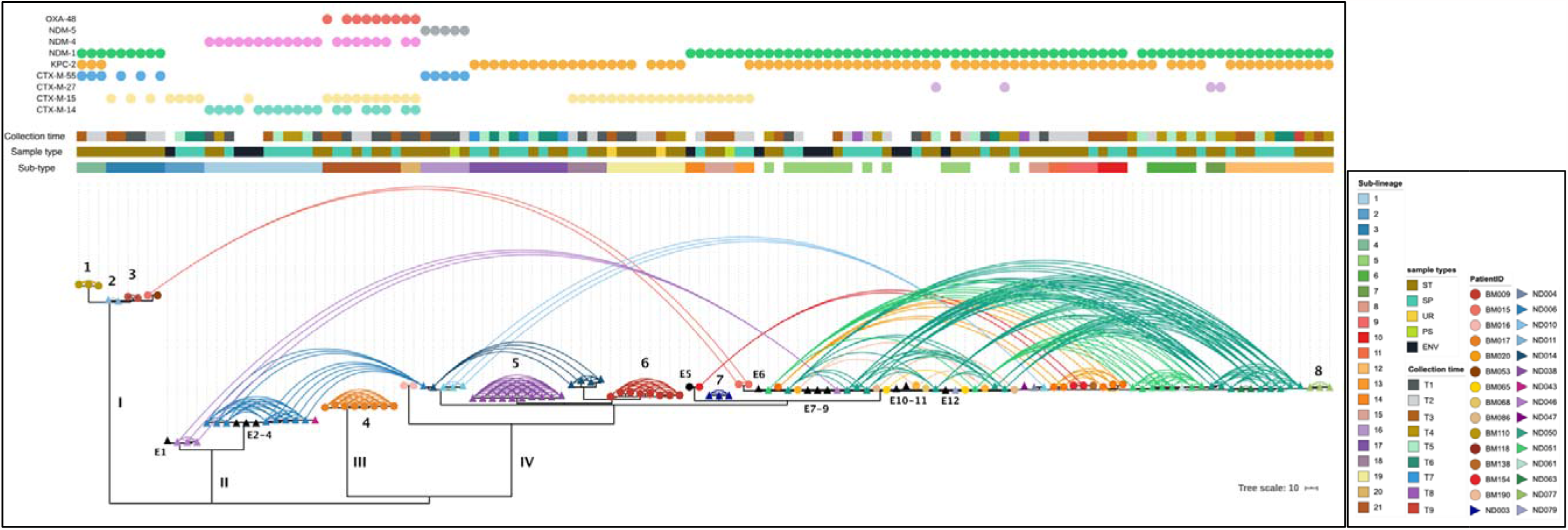
A maximum likelihood phylogeny and patient network of *K. pneumoniae* ST15 isolates from BMH and NHTD. Hospital is denoted by circles or triangles on branch tips for BMH or NHTD, respectively, coloured by patient (see key) or environmental samples appear as black circles or triangles. A pairwise linkage network between every sample from each individual patient is shown as arks, coloured as per patient. rPinecone sub-types, sample types and collection times are shown above. Unique colour codes for each sub-lineage are shown in legend. The presence of an ESBL- or carbapenemase-encoding gene(s) is indicated by coloured circles, top.

Secondly, whilst the majority of patient ND006 (ST15-1) and ND014 (ST15-18) isolates belonged to a single subtype, both also carried ST15-16 subtype isolates highly similar (median 4, IQR1-IQR3 2-11 SNP difference) to those first seen in patient ND010 two weeks earlier providing strong evidence for nosocomial transmission (Figures 2 & 3; Pattern 2 – ‘within-hospital transmission’). Whilst isolates identical to ST15-16 were not identified in environmental samples, ST15-1 isolates (1-5 SNP difference) were widely distributed across the ICU, including the bed spaces of patients ND014 and ND047, and in a stool sample from patient ND043 (Figures 2 & 3). These four patients were admitted to adjacent rooms in NHTD, shared the same basic facilities, and overlapping admission times (Figure 3).

**Figure 3:**
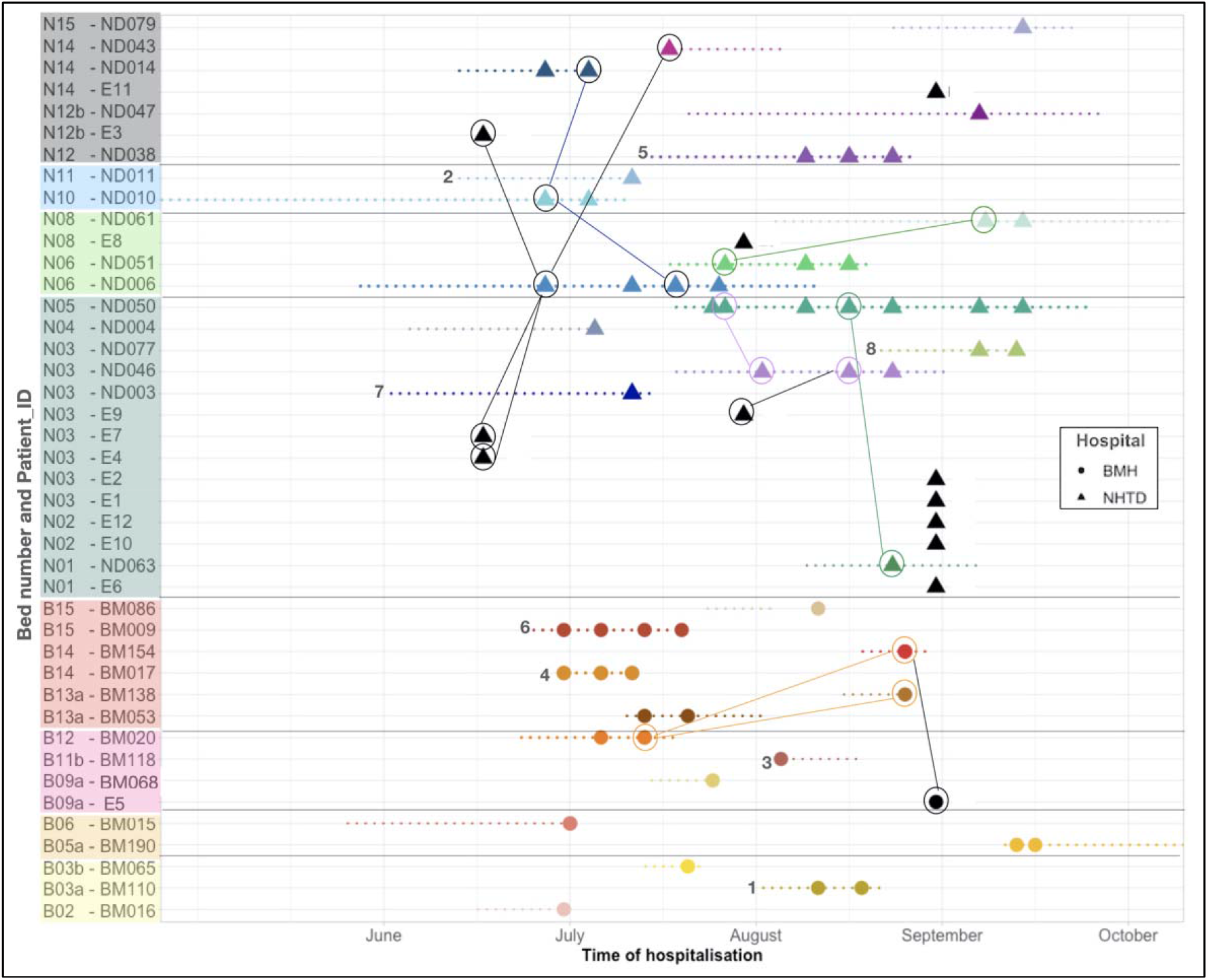
Epidemiological timeline of patients carrying *K. pneumoniae* ST15 at BMH and NHTD. Colour blocks include patients who stayed in the same room in the ICU wards, four from BMH (lower part) and four from NHTD (upper part). Each sample’s collection time (on admission, on discharge and weekly if patients stayed longer than 7 days in ICUs) within a patient is indicated by a single circle (BMH) or triangle (NHTD). Dotted lines represent the length of stay per patient. The links showed the connection between patients, or patients and the environment, that had the different isolates first before the subsequent patient was seen to have that strain.

Thirdly, 61 isolates taken from ten patients each from BMH and NHTD belonged to a branch comprised of nine subtypes, ST15-5 to ST15-13. These subtypes were differentiated by a median of 7 SNPs regardless of the hospital (e.g. ND051 median 6 SNPs, range 1-11 SNPs) similar to that seen within a single patient described by pattern one (Figure 2; Supplementary Figure 5A; Pattern 3 – ‘multi-hospital/community’). Although there is some evidence for Nosocomial transmission from patients isolates ND050, ND051 and BM020 with those from other patients with overlapping admissions (Figure 2 & 3; Supplementary Figure 5B), all patients carrying these subtypes shared a pool of highly related isolates equally distributed across both hospitals, with no hospital-specific phylogenetic signal.

Whilst there were fewer samples belonging to ST11, ST16, and ST656, we found some evidence that these three patterns of patient infection were reproduced in other STs (Supplementary Figure 6). Most isolates from patients carrying ST11 or ST656 belonged to a single lineage also shared across both hospitals. For ST11, isolates from both hospitals were differentiated by zero pairwise SNP differences. The phylogeny of ST16 was the exception of showing a more complex distribution within and between the two hospitals (Supplementary Figure 6).

### The emergence of *bla*_KPC-2_ carrying *K. pneumoniae* in Hanoi

None of the three patterns described above could be linked to the presence/absence of known accessory virulence genes that may explain the different patterns of transmission and carriage, for example those associated with invasive disease, the exception being *ybt* encoding yersiniabactin which is common across all STs (Supplementary Figure 1), or the capsular polysaccharide loci e.g. K-1 or K-2 associated with hypervirulent strains emerging in South East Asia [7] (Supplementary Figure 1, Supplementary Data Sheet 1). However, as noted above, >50% of the observed carbapenem-resistance could be explained by the presence of *bla*_KPC-2._ This contrasted previous reports showing *bla*_KPC-2-type_ was not detected in Southeast Asian isolates [7]. Here *bla*_KPC-2_ was found in 63.36% (n=83) ST15, 94.00% (n= 47) of ST11 and 91.00% (n= 40) of ST656 isolates. In addition, unlike previous studies showing most *K. pneumoniae* harboured either *bla*_KPC-2_ or *bla*_NDM-1_, of the 83 ST15 *K. pneumoniae* carrying *bla*_KPC-2_ reported here, 61 also carried *bla*_NDM-1_. It was also clear that whilst both genes were broadly distributed across *K. pneumoniae* sequenced here, only ST15 carried both concurrently (Supplementary Figure 1).

To understand the global relevance of this observation, we constructed a phylogeny of all available ST15 genomes, including the 131 genomes sequenced here, 33 published genomes from the Hanoi Vietnam National Children’s Hospital (VNCH) collected in 2015 [13], and 245 from globally dispersed isolates (Supplementary Data Sheet 3; Supplementary Figure 7). Of the Hanoi 2017 isolates, 98 fell on a single branch, thus far unique to Hanoi; and the remaining 33 formed three clades distributed across the ST15 global phylogeny. Interestingly, all isolates from VNCH formed a sister clade with the main Hanoi 2017/18 clade IV isolates from the current study. These data also showed that within ST15, apart from 3 isolates from Clade I (K-type 112; Figure 2; Supplementary Figure 1) all other ST15 isolates carrying *bla*_KPC-2_ and *bla*_NDM-1_ were described by Pattern 3 in BMH and NHTD or linked to VNCH.

We identified a putative IncN plasmid (we denoted pMHP-KPC2) in the ST15 *bla*_KPC-2_ positive assemblies. pMHP-KPC2 shared >99% nucleotide identity (covering >92 % of the sequence length) with pEC_224: an *E. coli* IncN *bla*_KPC-2_ plasmid taken from a patient involved in the Study for Monitoring of Antimicrobial Resistance Trends in Hanoi in 2010 (accession No. NZ_CP018945.1; Supplementary Figure 8, Supplementary Table 3). In addition to *bla*_KPC-2_ plasmid SR6656629, isolated from VNCH *K. pneumoniae* 2015 isolates, which appears to be a truncated form of pMHP-KPC2 lacking mobility and transfer functions (Supplementary Figure 9). Either pMHP-KPC2 or SR6656629 were present and conserved (98.46% average coverage and 96.38% average identity) in all *bla*_*K*PC-2_ containing isolates seen here, regardless STs, except for five isolates of ST11 from patient ND045 where the pMHP-KPC2 *bla*_KPC-2_ gene cassette was chromosomally located (Supplementary Figure 9).

Whilst *bla*_NDM-1_ was not co-located with *bla*_KPC-2_ on pMHP-KPC2, all *bla*_NDM-1_ containing isolates from Hanoi carried a 26 kb conserved core sequence (95.05% average coverage and 98.68% average identity; Supplementary Figure 10) bordered by multiple mobile elements, including a truncated copy of Tn125 and *qnrS1* genes, conferring quinolone resistance.

## DISCUSSION

In Vietnam, antimicrobials are widely available without prescription, and there is a culture of self-medication prior to seeking medical attention or hospitalisation [14, 15]. Within the clinical setting, third-generation cephalosporins are the most commonly used antimicrobials for community- and hospital-acquired infections, followed by fluoroquinolones and carbapenems [16]. However, within the ICU setting, carbapenems were used most, accounting for 22.9% of all antimicrobials administered [16]. We found that 90% of the ESBL-P *K. pneumoniae* carriage strains isolated here were also resistant to carbapenems, with the most common agents being meropenem and ertapenem. Importantly, although piperacillin-tazobactam has been considered as an alternative to carbapenems for patients in ICU [17], here 96.63% of the isolates collected were seen to be phenotypically resistant. However, a considerably lower proportion of the ESBL-P or CR isolates remained susceptible to amikacin (57.70%), gentamicin (49.01%), and tigecycline (35.01%), thus offering some limited therapeutic options.

From the genomic data we identified 22 *K. pneumoniae* STs of which 88% belonged to 5 STs: ST11, ST15, ST16, ST147, ST656. Of which ST11, ST15 and ST147 are known as MDR lineages linked to numerous outbreaks in Europe and Asia [18-20]. ST16 is of increasing concern in Asia [21], but ST656 has only previously been found in sporadic cases in the Philippines and China [22, 23]. Within these STs, *bla*_CTX-M-15_ was the most common ESBL gene, consistent with previous reports [24], and *bla*_KPC-2_ was the most common carbapenemase gene, present in over 50% of all isolates sequenced here. The latter contrast previous data from bloodstream isolates collected in South and South East Asia (collected between 2010-2017; including 89 isolates from Vietnam) [7] where no KPC class carbapenemase genes were observed. In addition, the overall number of STs identified from blood was greater than seen here: 120 STs, of which ∼30% belonged to four STs [7], with the only overlap being ST15 which was the most common ST in both studies [7]. Consistent with sample type, ST23 - considered to be an emerging Asian hypervirulent lineage and the second most common ST seen in blood [7], was absent from the BMH and NHTD samples analysed here.

The increase in the prevalence of carbapenem resistance, and particularly KPC class genes, seen here is important and reflects the overall increase in resistance to carbapenems seen in South East Asia. KPC-producing *K. pneumoniae* are difficult to detect using routine tests and so are linked to treatment delays and high mortality rates [25]. *bla*_*K*PC-2_ was also an important factor in the global spread of ST258, one of the most successful high-risk *K. pneumoniae* STs [1]. Unlike ST258, where *bla*_*K*PC-2_ is linked to the IncFII plasmid pKpQIL [26, 27], here *bla*_*K*PC-2_ was located on self-mobilisable IncN plasmids, seen first in *E. coli* isolated in Hanoi in 2010 (pEC_224), then subsequently in all isolates from an outbreak reported in 2015 of resistant *K. pneumoniae* in a Hanoi Paediatric hospital (SR6656629; VNCH) and now here in isolates collected in 2017/18 (pMHP-KPC2).

We also found that 61/184 of the *bla*_*K*PC-2_ isolates sequenced here had subsequently acquired *bla*_NDM-1_. Whilst the dual *bla*_*K*PC-2_/*bla*_NDM-1_ genotype has been seen previously, including in one of the 2015 outbreak isolates, until now it has been rare. This was previously explained by the implied fitness cost of carrying two functionally redundant genes [13, 28]. These genes represent both classes of carbapenemases (the serine and metallo-beta lactamases), so it is important to understand if the high prevalence and persistence of this dual resistance in ST15 has emerged locally and moved through these hospitals/communities where there is high carbapenem usage. This phenomenon may pose a serious future problem in these healthcare settings.

BMH and NHTD do not share staff or equipment, and none of the patients sampled here was transferred between these two hospitals. Longitudinal analysis of *K. pneumoniae* ST15 carriage isolates showed three patterns of infection evident in both hospital ICUs: 26.67% carried the same *K. pneumoniae* subtype from their first sample in or around admission until discharge from ICU. Compared to all other isolates sequenced these isolates were genetically unique to that individual patient, suggestive of persistent carriage and importation from home or the previous referral hospital/ ward (Pattern 1). Furthermore, a minority of patient isolates could be linked to intra-hospital transmission (four patients with 26 isolates; Pattern 2).

However, the most commonly observed pattern of carriage (Pattern 3) was for isolates belonging to a restricted branch of ST15 describing subtypes present across both ICUs and varying by <11 SNPs, regardless of hospital ICU. Here the high clonality of the ST15 isolates, relative to other ST15 sublineages, is taken to indicate a common source of transmission. Whilst we found some evidence of nosocomial transmission within BMH and NHTD ICUs, our data suggest that the majority of ESBL-positive and/or carbapenem-resistant *K. pneumoniae* were distributed widely across the different levels of hospital care. The patterns of transmission we observed here can be explained by these carriage isolates being part of widely distributed, almost clonal, endemic populations circulating in the referral hospitals and/or the communities that feed both BMH and NHTD directly.

These three patterns of transmission and carriage were reflected in most other STs collected. They are important because of the mounting evidence that hospital-associated bloodstream infections originate from the isolates carried in a patient’s own microbiome [29], and because of the current reliance on carbapenems for the treatment of Gram-negative bacteraemia. A limitation of this study was that whilst it is possible to track referrals into a hospital the hospital for which the patient is being referred is not always well documented. This has improved recently with the institution of an electronic patient record system. However, a key strength of this study is the use of multi-site screening of patients across two critical care settings. Focusing surveillance activities on a single hospital makes sense if the emergence of important pathogens like *K. pneumoniae* occurs within that hospital, a notion that is strongly held. By linking surveillance activities across hospitals, we used these hospitals as lenses to view both the carriage of clinically relevant *K. pneumoniae* within and between patients and hospitals, showing that most of the isolates showed no hospital-specific profile (Supplementary Figure 1).

Our findings emphasise that it is insufficient to simply study a single hospital, in fact this may even be misleading. Further, solely studying the hospital referral network may not provide enough comprehensive information. What is needed, especially where there is unrestricted use of antimicrobials, is a truly integrated surveillance system which considers the greater community as well as individual wards, hospitals and the referral system. In doing so, it enables us to rationally target measures designed to reduce the emergence of antimicrobial resistance with implications for infection control as well as health and safety interventions at all hospital levels.

## Data Availability

All raw and assembled Illumina WGS data are available on the European Nucleotide Archive under the Bio-project PRJEB29424. A list of individual sample accession numbers is included in Supplementary Data Sheet 1.

## ETHICAL STATEMENT

Written informed consent for study participation was obtained from all study participants or their next of kin. The study protocol was reviewed and approved by the Institutional Review Boards of the National Hospital for Tropical Diseases (reference: 241/NDTW-TCCB) and Bach Mai Hospital (reference: 133/CV-BM), Hanoi Vietnam, and by the University of Cambridge Human and Biology Research Ethics Committee, United Kingdom (reference HBREC.2017.09). The study was registered with the ISRCTN registry (reference: ISRCTN18160232).

## ACKNOWLEDGEMENTS

The authors would like to thank all of the patients who participated in this study. We also acknowledge the clinical and laboratory staff from the National Hospital for Tropical Diseases and Bach Mai Hospital for their assistance with this study. Finally, we acknowledge the support of Christoph Puethe and the Pathogen Informatics team at the Wellcome Sanger Institute. Last but not least, we want to show our gratitude to the sequencing team at the Wellcome Sanger Institute for generating the whole genome sequencing data.

