## Supplementary Material for "Evidence of widespread endemic populations of highly multidrug-resistant *Klebsiella pneumoniae* seen concurrently through the lens of two hospital intensive care units in Vietnam"

### SUPPLEMENTARY METHODS

#### Whole genome sequencing (WGS) and Quality Control (QC) analysis

Genomic DNA was extracted using QIACube biorobots and the QIAamp 96 DNA QIAcube HT and sequenced on an Illumina HiSeq X10 with 125bp pair-ended reads. Read quality was checked by FastQC v0.11.4 [1], and contaminated samples were identified using Kraken v0.10.6 [2] and Bracken v2.2 [3] (samples with >30% non-*K. pneumoniae* reads were excluded). Assemblies were generated using SPAdes v.3.11 [4] with the genome size distribution checked using QUAST v4.6 [5], genomes containing > 300 contigs were excluded. We screened our assemblies using CheckM v1.0.5 [6] for contamination and genome completeness, retaining assemblies with contamination ≤ 5% and completeness ≥ 99%. Annotation was performed using PROKKA v1.11 [7].

#### Genotypic features and phylogenetic analysis

Kleborate v0.3.0 [8, 9] was used to identify the multi-locus sequence types (MLST), antimicrobial resistance and virulence genes, with a blastp ≥95% identity cut off. K-antigen and O-antigen were determined using Kaptive [10, 11], with ≥90% coverage thresholds. Plasmid types were identified using ARIBA [85] with a ≥ 90% blastp identity cut off against the PlasmidFinder database [12]. The core and accessory genomes were determined using Roary [13] (≥ 95% blastp identity cut off). Single nucleotide polymorphisms (SNPs) were extracted from the core-gene alignment using snp-sites [14]. Maximum-likelihood phylogenetic trees were constructed from SNP-only alignments using IQ-TREE v1.6.5 [15], adding the number of constant sites with '-fconst' option, and using GTR+F+I+G4 model with 1000 ultrafast bootstrap replicates [16]. Phylogenetic trees and complex antimicrobial resistance profiles were visualised using Phandango [17].

#### Phylogenetic analysis of *K. pneumoniae* ST15

131 *K. pneumoniae* ST15 genomes from this study were combined with 278 publicly available *K. pneumoniae* ST15 genomes to provide global context. We used MASH [18] to identify the closest matching reference sequence (*K. pneumoniae* PMK1 strain, NCBI accession no: CP008929) and mapped reads to this reference using BWA-MEM v1.2 [19], followed by indel realignment using GATK v3.4.46 IndelRealigner [20], deduplication with Picard MarkDuplicates v1.127

(http://broadinstitute.github.io/picard/), and variant calling and consensus pseudosequence generation using samtools v1.2 [21] and bcftools v1.2[21]. A minimum of 8 supporting reads (3 per strand) and a variant frequency/mapping quality cut-off of 0.8 were used to call variants. Sites not meeting these criteria were masked to 'N' in the pseudosequence. The pairwise SNP distances between *K. pneumoniae* ST15 isolates from NHTD and BMH were obtained using pairsnp (https://github.com/gtonkinhill/pairsnp). Recombination was identified and masked using Gubbins v 1.4.10 [22]. The recombination-free phylogenetic tree was constructed using IQ-TREE v1.6.5 [15], inputting the number of constant sites with '-fconst' option, and using a TVM+F+I model and 1000 ultrafast bootstrap replicates [16]. Pyjar (https://github.com/simonrharris/pyjar) was used to perform joint ancestral reconstruction of substitutions [23], and sub-lineage clusters were inferred using rPinecone [24] with a 10 SNP threshold. Pairwise patient linkage networks were permuted in R v4.0.2 [25], and patient timelines and interactions were plotted in R using ggplot2 v3.3.3 [26]. iTOL [27] was used to visualise phylogenies with metadata.

**Plasmid comparisons**

Circularised *bla*$_{KPC-2}$- carrying plasmids were annotated and mapped against the reference using BRIG [28]. The comparison of synteny between two or more sequences were visualised using the Artemis Comparison Tool v13.0.0 [29]. The core sequence of *bla*$_{NDM-1}$- containing contig was constructed using Genious v11.1.5 [30].

**Statistical analysis:**

Bayesian Multinomial and Poisson regression models were fitted to the ST and within hospital SNP data, respectively, to determine their association with the hospital from which isolates were obtained, using rstan [25]. To correct for multiple sampling, a maximum of one observation per patient for each unique ST-AMR profile was used in the multinomial model, while sampling time-differences between each pair of isolates were adjusted for in the Poisson model. We quantified concordance of AMR genes and phenotype as the proportion of isolates that had both the gene and phenotype to the number of isolates with the gene.

### SUPPLENTARY RESULTS

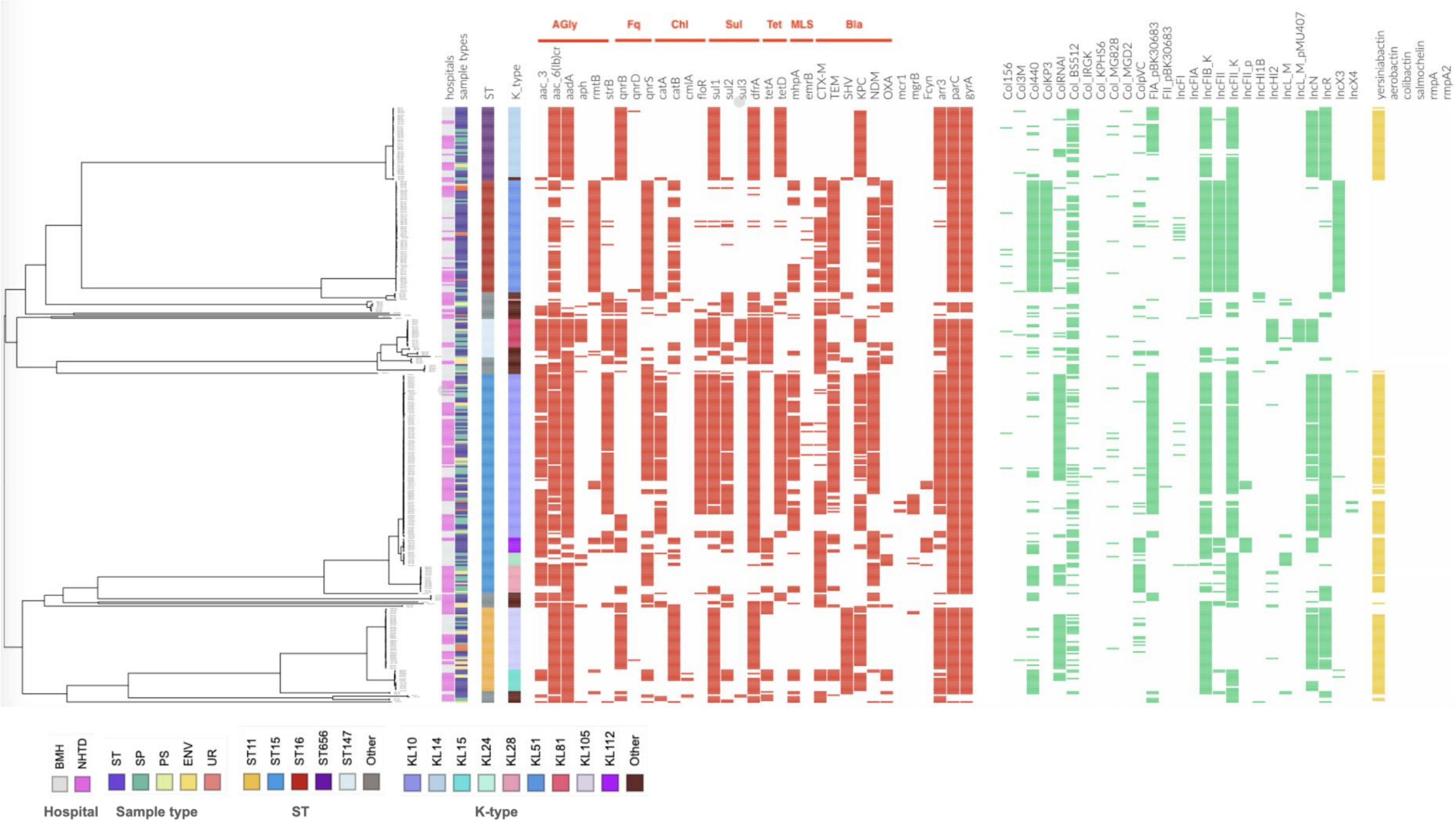


**Supplementary Figure 1: Antimicrobial resistance profile of 357 *K. pneumoniae* isolates collected from Bach Mai Hospital (BMH) and the National Hospital for Tropical Diseases (NHTD)**

The guide tree is based on core-snp alignment on Roary. The colour blocks show the diversity of sample type, sequence type, capsule type, and the presence/absence of each antimicrobial resistance genes, plasmid replicons and virulence genes.

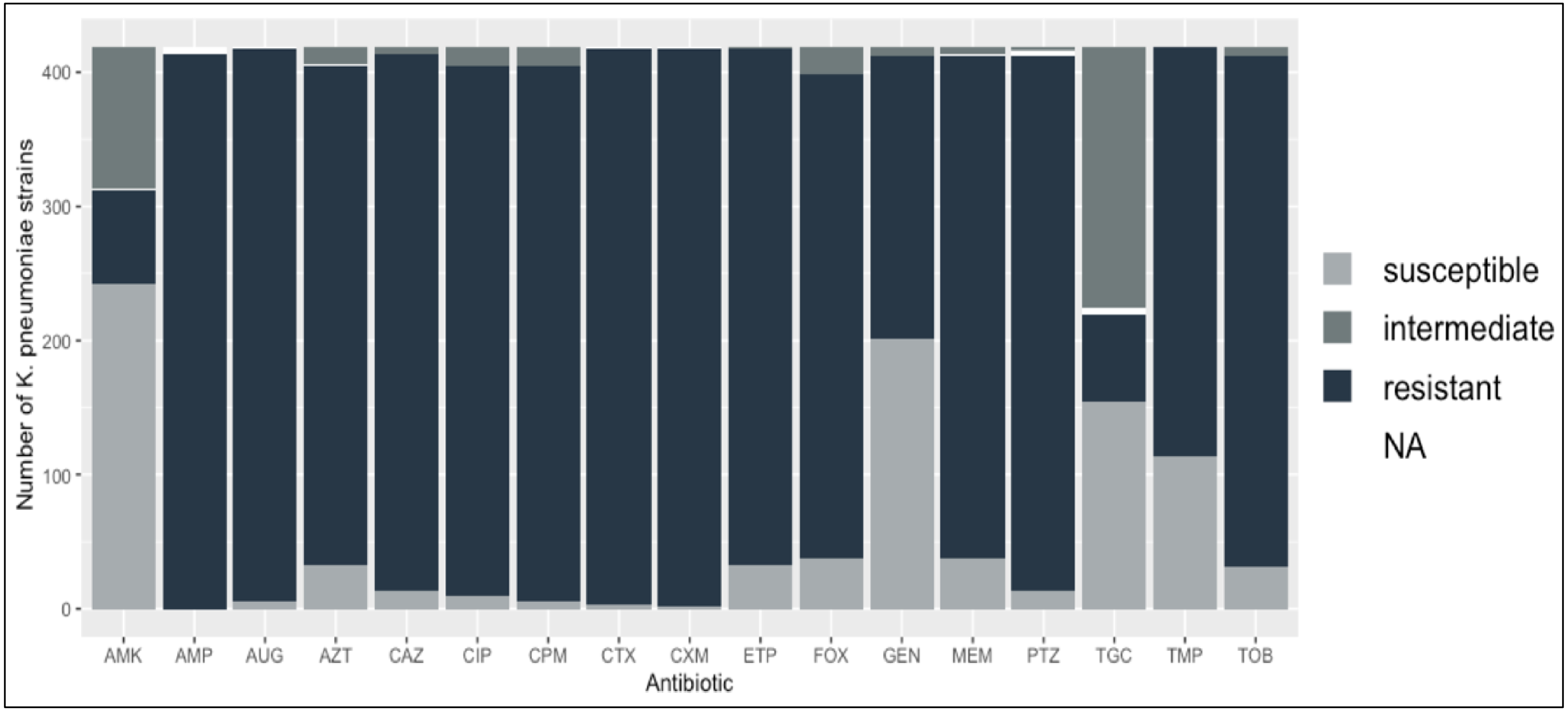


**Supplementary Figure 2: Vitek-2 antimicrobial susceptibility profiles of *K. pneumoniae* isolates**

Different shades of grey represent different level of antibiotic susceptibility. White blocks indicate samples with missing results. Antimicrobial abbreviation: amikacin (AMK), ampicillin (AMP), amoxicillin/clavulanic acid (AUG), aztreonam (AZT), ceftazidime (CAZ), ciprofloxacin (CIP), cefepime (CPM), cefotaxime (CTX), cefuroxime (CXM), ertapenem (ETP), cefoxitin (FOX), gentamicin (GEN), meropenem (MEM), piperacillin-tazobactam (PTZ), tigecycline (TGC), trimethoprim (TMP), tobramycin (TOB).

**Supplementary Figure 3: Carbapenemase and ESBL gene-phenotype concordance.**

Panels a-c show the concordance of six carbapenemase encoding genes (*bla*$_{KPC-2}$, *bla*$_{NDM-1}$, *bla*$_{NDM-4}$, *bla*$_{NDM-5}$, *bla*$_{OXA-48}$ and *bla*$_{OXA-181}$) identified in the genomes of the *K. pneumoniae* isolates from two hospitals in Hanoi and resistance to three carbapenems (Meropenem, Ertapenem, and Aztreonam).

Panels d-h show concordance between six ESBL encoding genes (*bla*$_{CTM-15}$, *bla*$_{CTXM-14}$, *bla*$_{CTX-M-3}$, *bla*$_{CTX-M-27}$, *bla*$_{CTX-M-55}$, *bla*$_{SHV-12}$) identified in Hanoi *K. pneumoniae* genomes and resistance to third generation cephalosporins (Cefepime, Cefotaxime, Cefoxitin, Ceftazidime, and Cefuroxime). Concordance of AMR genes and phenotype as the proportion of isolates that had both the gene and phenotype to the number of isolates with the gene.

Panel i shows the number of genomes that had the gene to contextualise the proportions in panels a-h.

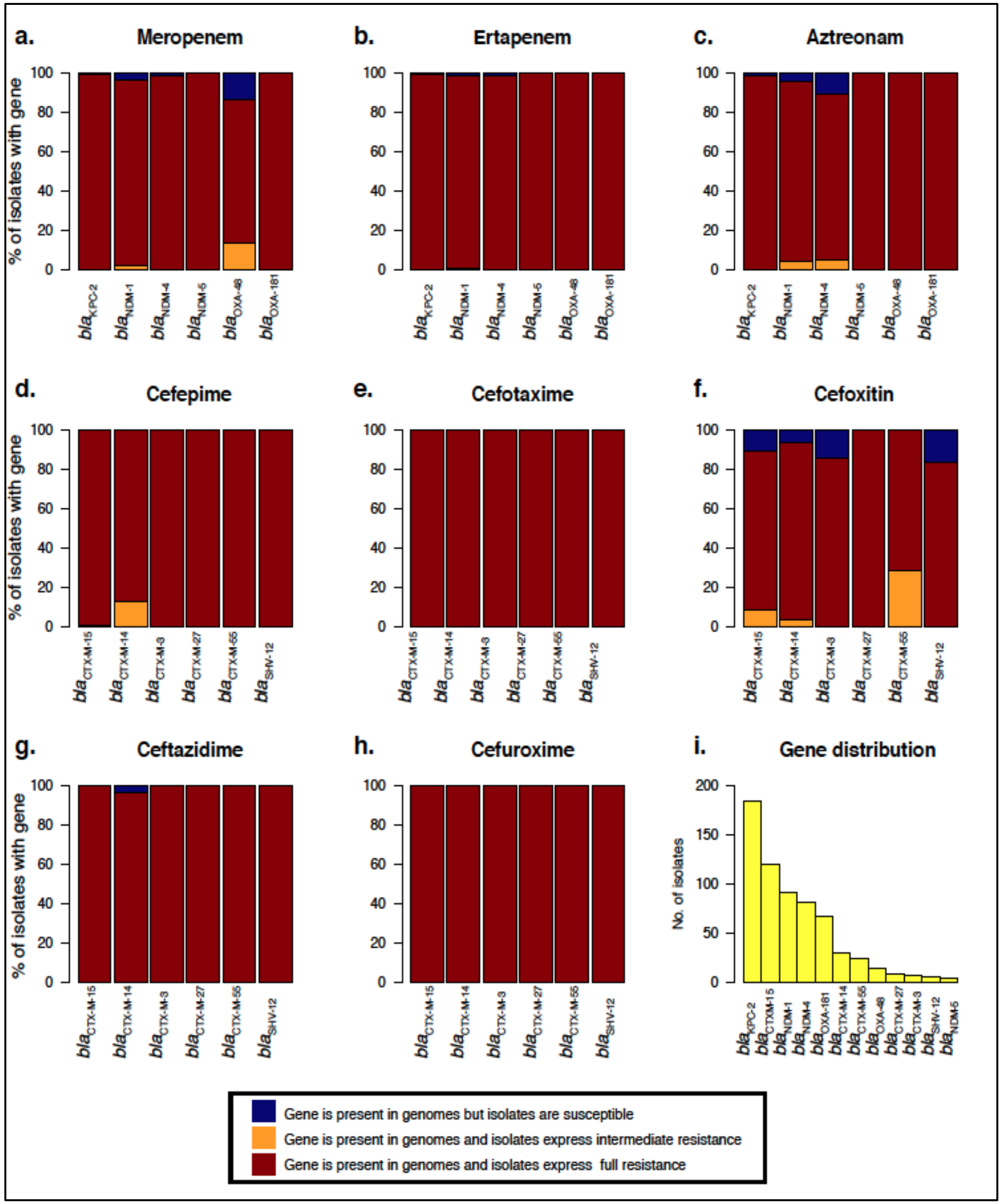

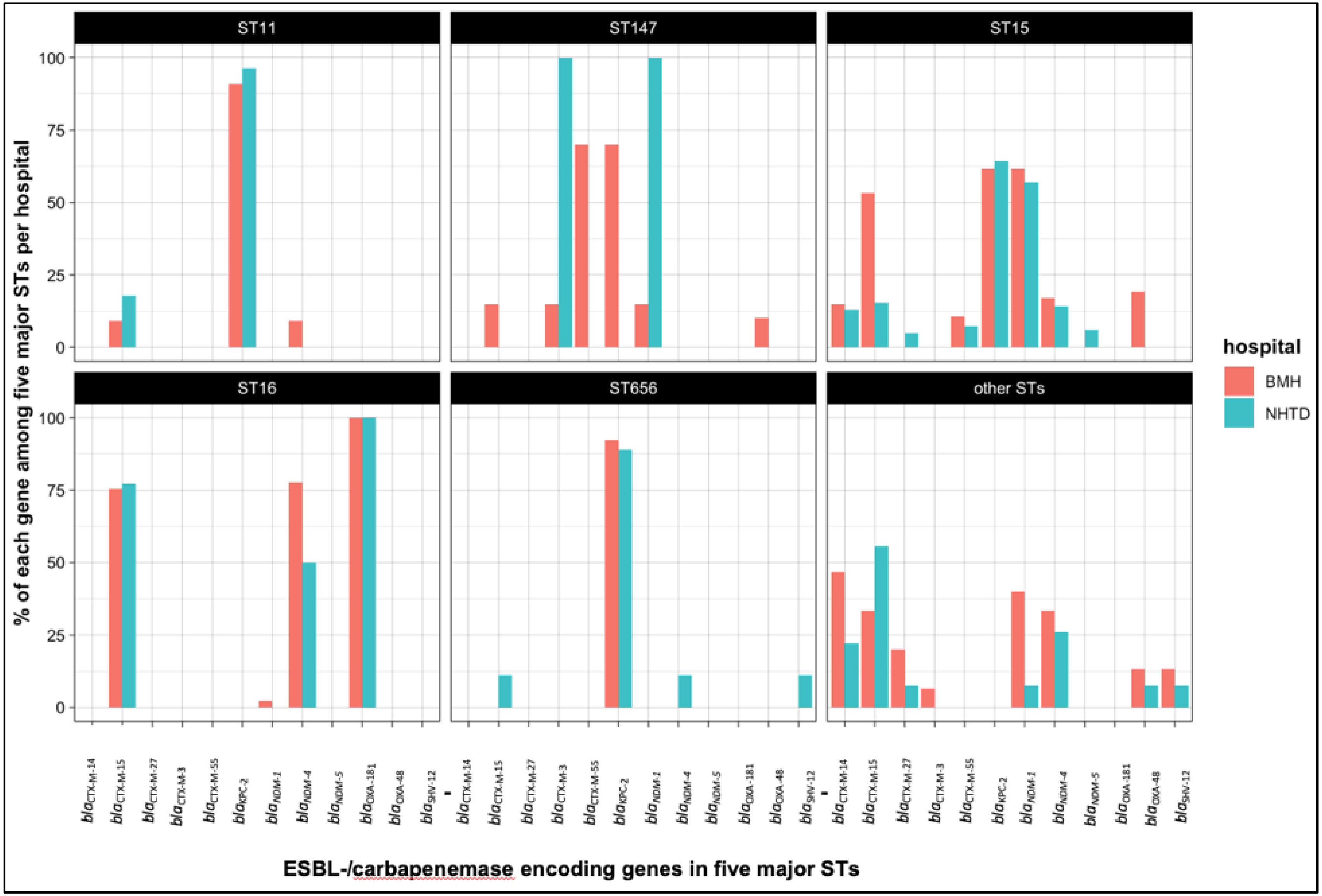


**Supplementary Figure 4: The prevalence of ESBL-/carbapenemase encoding genes in five major STs of *K. pneumoniae* isolates from Bach Mai Hospital (BMH) and the National Hospital for Tropical Diseases (NHTD). Bar plots show the frequency of each ESBL-/carbapenemase encoding genes in the five major *K. pneumoniae* STs seen in this study (ST11, ST15, ST16, ST147, ST656).**

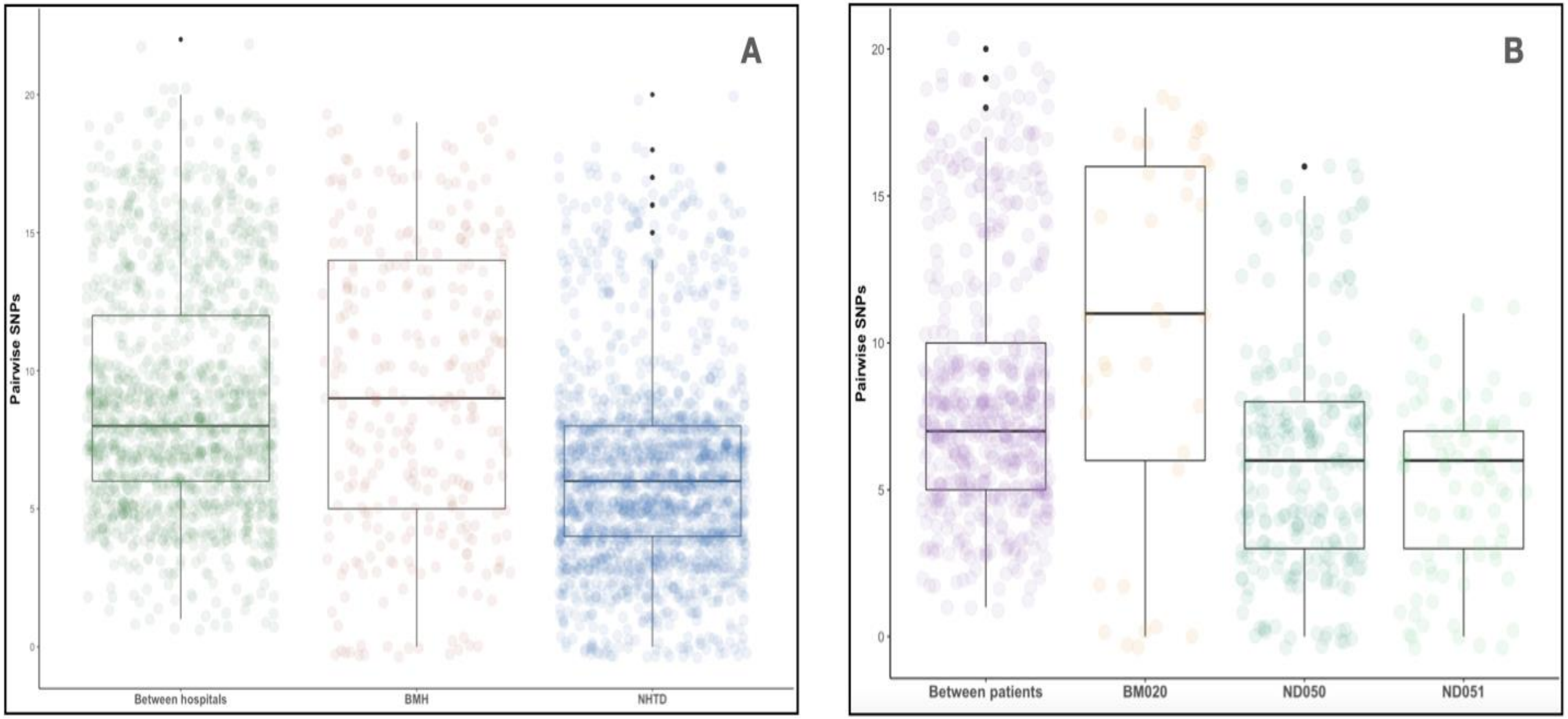


**Supplementary Figure 5: Pairwise distances of SNPs of *K. pneumoniae* ST15 isolates**

(A) of the high clonal cluster within BMH, within NHTD and between the two hospitals. There are 64 isolates in this cluster, 20 from BMH and 44 from NHTD. Each coloured dot represents the number of SNPs between two isolates in each group.

(B) of three patients with multiple samples belonged to the high conal cluster, in which, patient BM020 from BMH and patient ND050 and ND051 from NHTD.

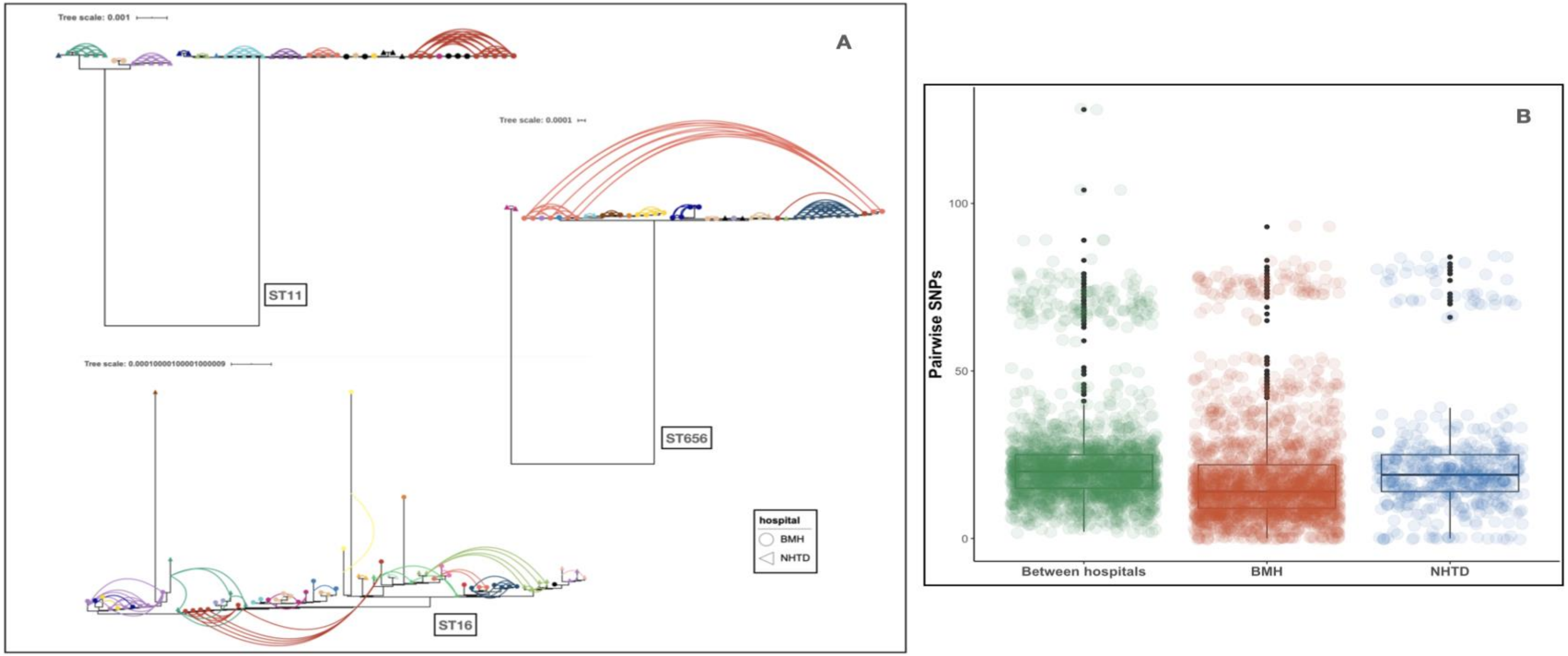


**Supplementary Figure 6: Phylogeny and network of patients carrying *K. pneumoniae* isolates within ST11, ST16 and ST656.**

(A): The guide tree was constructed based on multiple alignment mapping to the reference, then refined using a root-to-tip directional approach. The curved link was constructed for every sample pair from the same patients. The shape of tips represents a hospital, circle for BMH and triangle for NHTD. Colours are unique for each patient from BMH and NHTD. Environmental samples are coloured in black.

(B): Pairwise SNP distances between ST16 *K. pneumoniae* isolates within and between the two hospitals

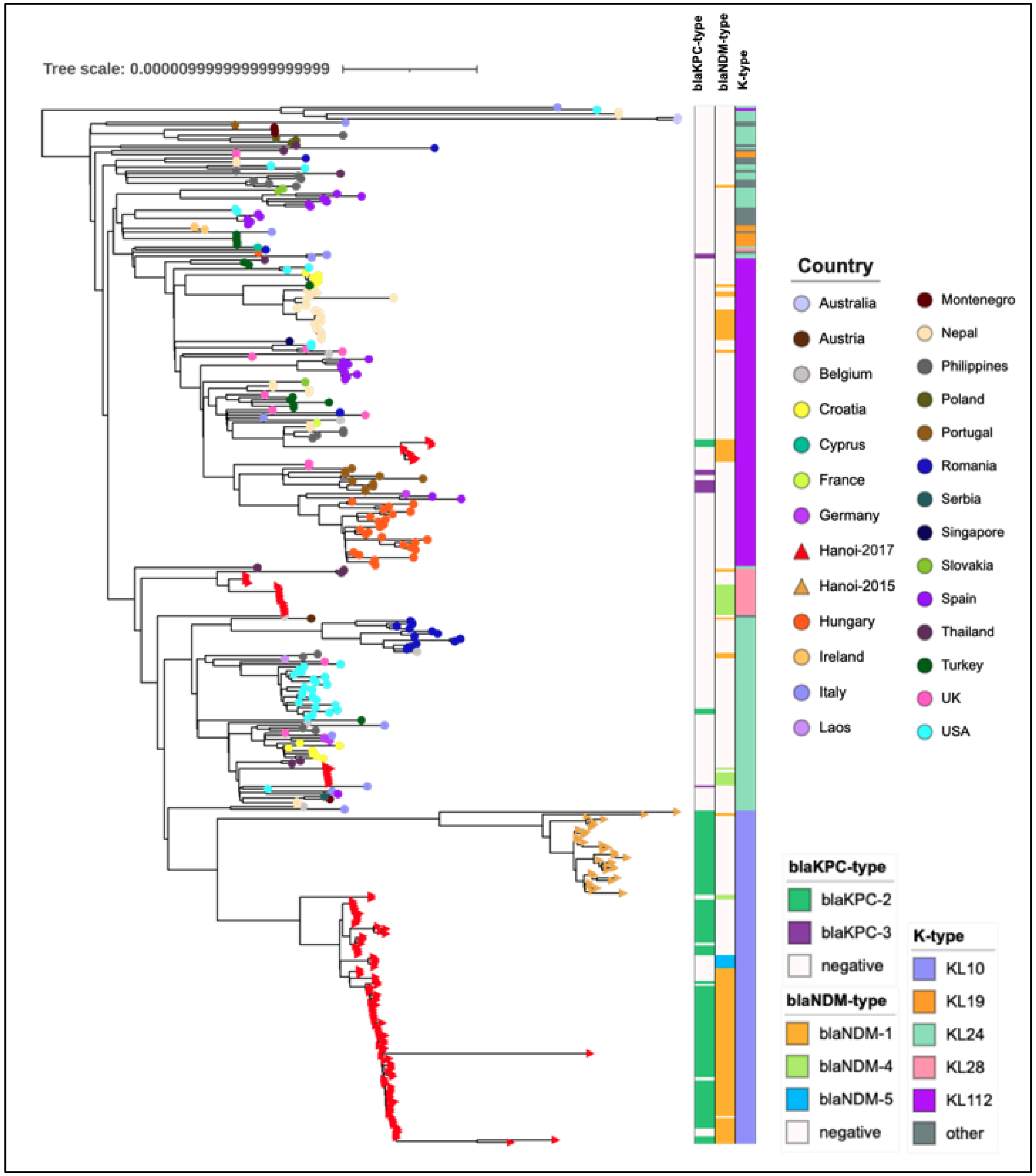


**Supplementary Figure 7:**

Maximum likelihood phylogeny of 131 ST15 *K. pneumoniae* isolates from Hanoi and 278 publicly available ST15 *K. pneumoniae* sequences.

Each coloured tip represents a country. Coloured strips indicate the presence and absence of $bla_{\text{KPC-type}}$ gene and $bla_{\text{NDM-type}}$ gene, and the distribution of the capsule K-types.

**Supplementary Figure 8: Genetic map of a conjugative *bla*$_{\text{KPC-2}}$ harboring IncN plasmid to the reference pEC224_IncN**

IncN plasmid has well conserved backbone. The backbone contains genes responsible for mobilization, and for replication and maintenance (annotated in black). The gene *bla*$_{\text{KPC-2}}$ (red arrow) is flanked by IS*Kpn27* and IS*Kpn6* (grey arrows). It is adjacent to a Tn3 transposon (grey) and Tn3-Derived Inverted-Repeat Miniature Elements, highlighted in orange.

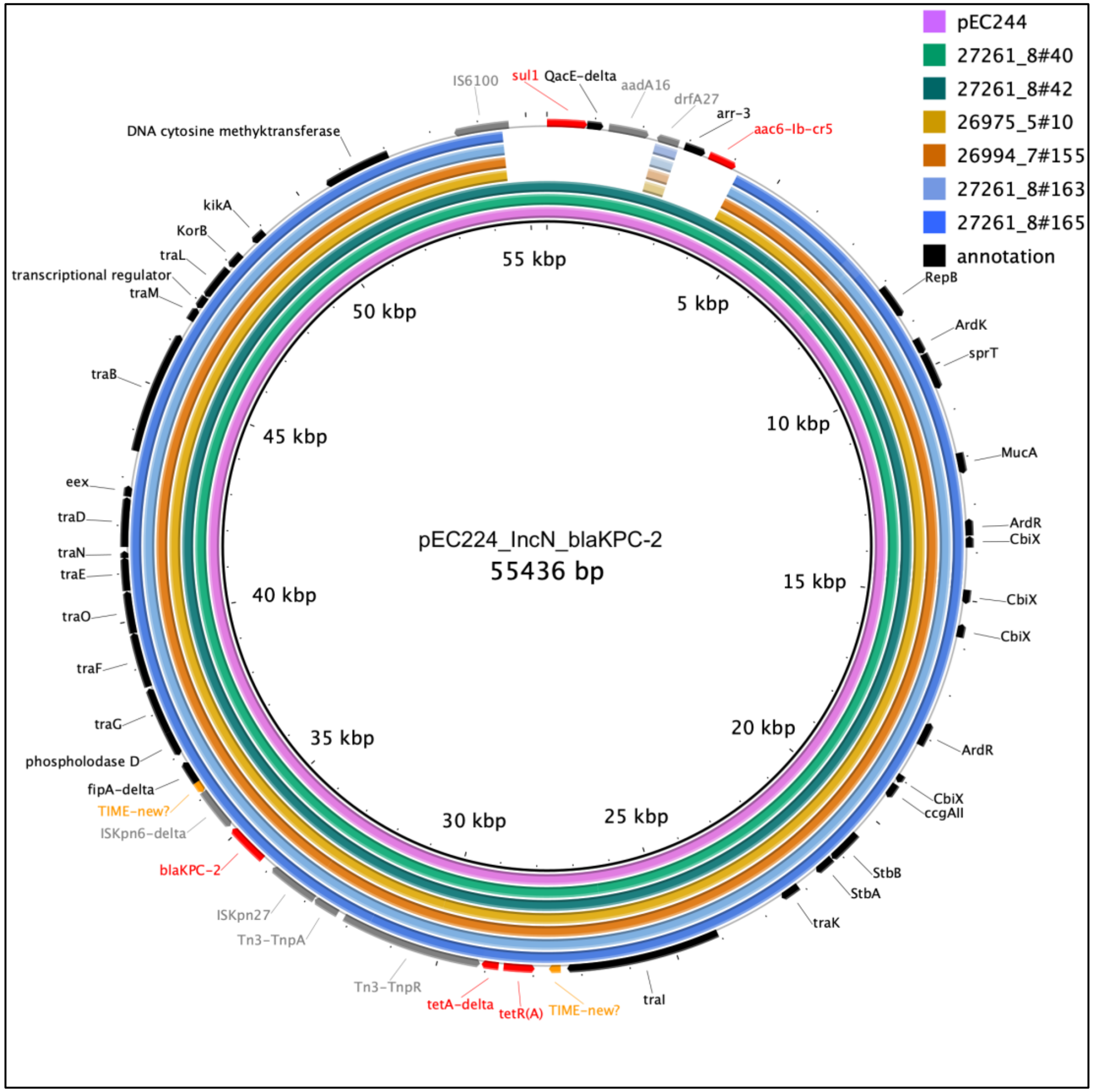

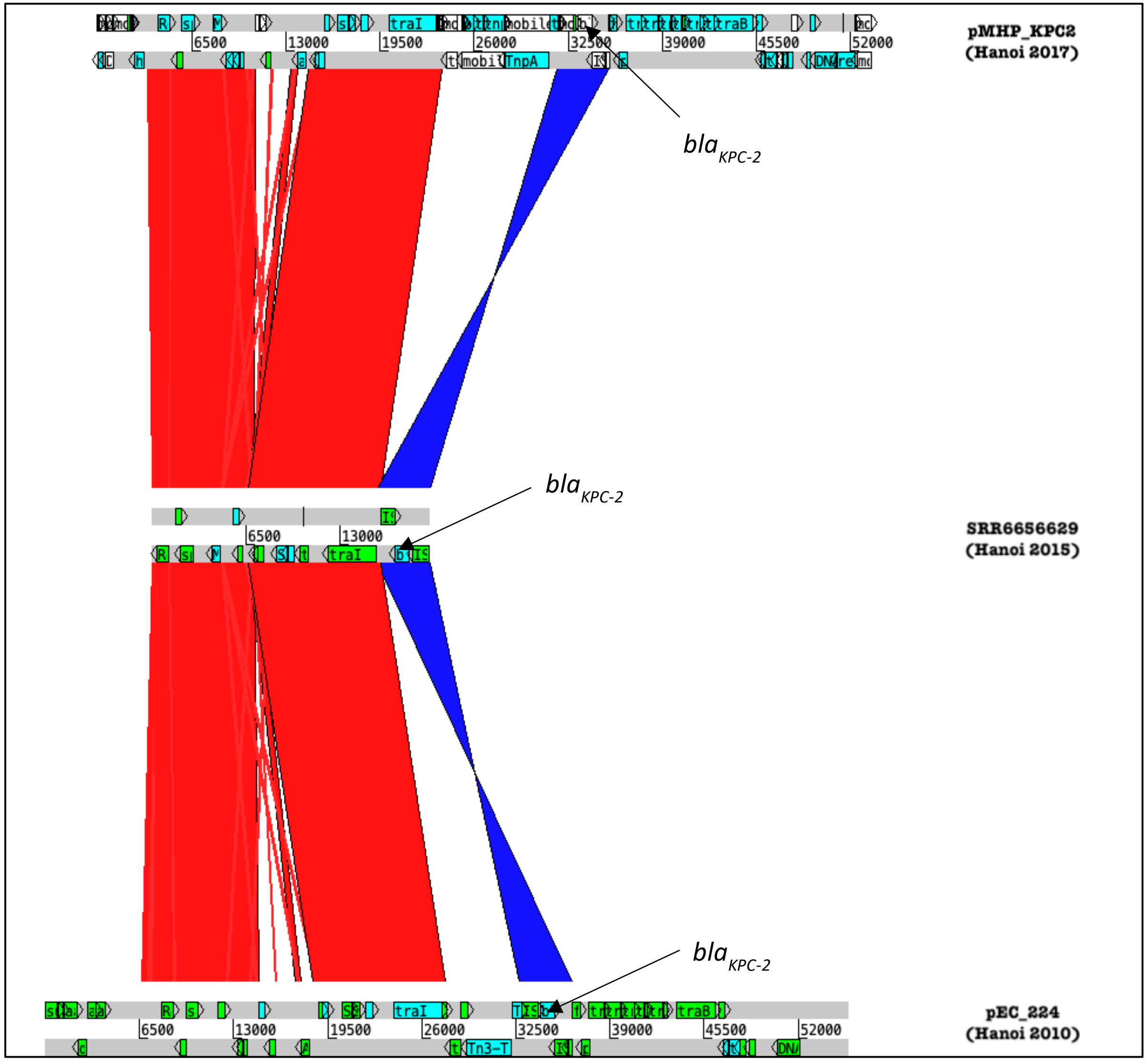


**Supplementary Figure 9: Structural similarity between two conjugative *bla*$_{KPC-2}$-harboring IncN plasmids and a *bla*$_{KPC-2}$-carrying contig.**

A comparison using the Artemis Comparison Tool (ACT) shows the high structural similarity between plasmid pMHP_KPC2 (circularised, isolated from *K. pneumoniae* in Hanoi 2017), plasmid pEC224 (circularised, isolated from *E.coli* in Hanoi 2010) and the contig SRR6656629 (isolated from *K. pneumoniae* in Hanoi 2015).

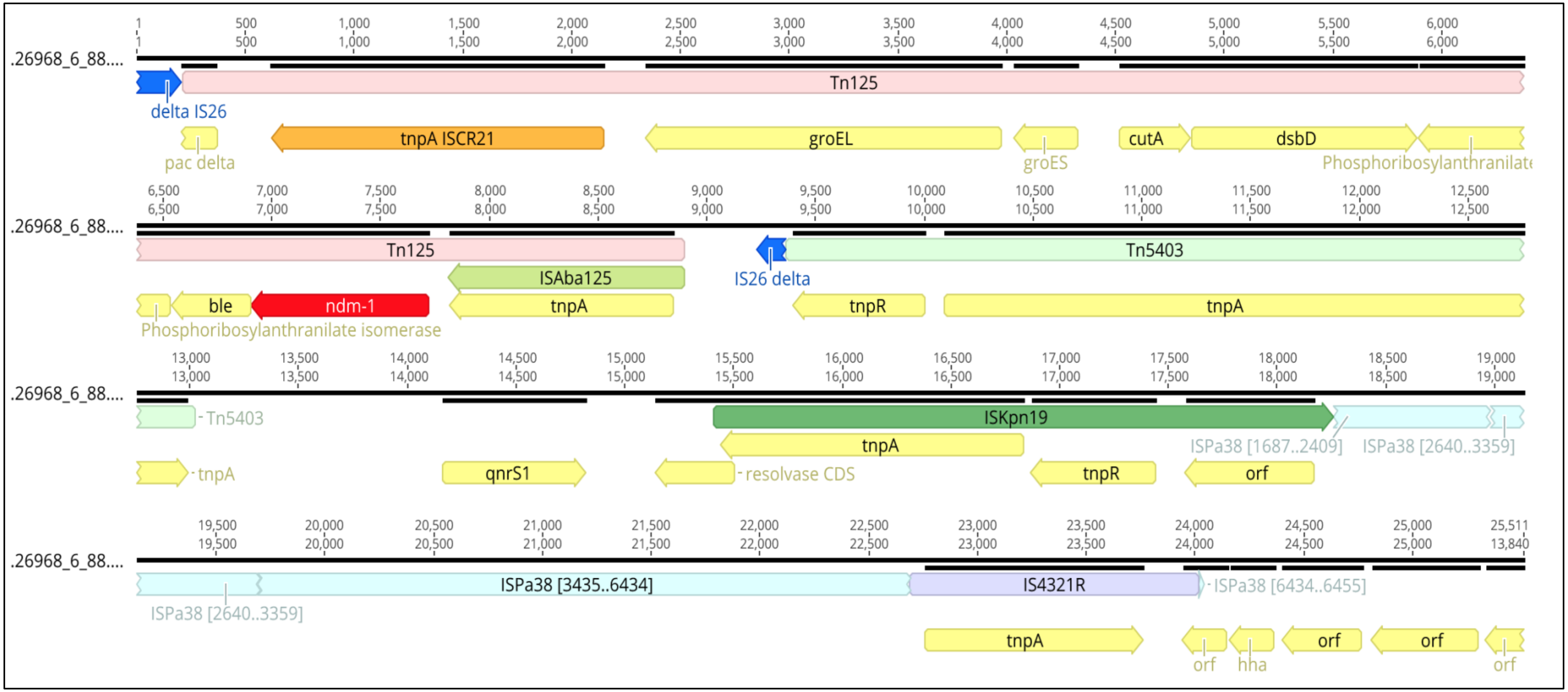


**Supplementary Figgure 10: The genetic structure of 26kb contig containing *bla*$_{NDM-1.}$**

The *bla*$_{NDM-1}$ was a part of the Tn*125* composite transposon which can be located on either chromosome or plasmid. The contig structure also shows the transposon Tn*5403* and the inserted sequence fragments IS*Kpn19* linked to the downstream of *bla*$_{NDM\_1}$

**Supplementary Table 1: Frequency of sequence types (STs) among 357 *Klebsiella pneumoniae* study isolates**

| Sequence type | No. of isolates | | Sequence type | No. of isolates |
|---|---|---|---|---|
| ST15 | 131 | | ST359 | 2 |
| ST16 | 67 | | ST395 | 2 |
| ST11 | 50 | | ST101 | 1 |
| ST656 | 44 | | ST199 | 1 |
| ST147 | 23 | | ST2938 | 1 |
| ST307 | 7 | | ST323 | 1 |
| ST20 | 5 | | ST337 | 1 |
| ST273 | 5 | | ST36 | 1 |
| ST327 | 4 | | ST685 | 1 |
| ST37 | 4 | | ST7 | 1 |
| ST392 | 4 | | ST709 | 1 |

**Supplementary Table 2: Prevalence of ESBL and carbapenamse-encoding genes among *K. pneumoniae* isolates**

| *bla*-genes | Number of isolates (%) | Number of STs (3 most common STs) | Gene variants (number of isolates) |
|---|---|---|---|
| ***bla*$_{CTX-M-like}$** | 193 (54.06%) | 19 (ST15, n= 64; ST16, n =51; ST147, n= 23) | CTX-M-15 (n= 120)<br>CTX-M-14 (n= 32)<br>CTX-M-55 (n= 25)<br>CTX-M-27 (n= 9)<br>CTX-M-3 (n=7) |
| ***bla*$_{SHV-like}$** | 6 (1.68%) | 3 (ST392, n= 3; ST656 n= 2; ST36, n=1) | SHV-12 (n= 6) |
| ***bla*$_{KPC-like}$** | 184 (51.54%) | 4 (ST15, n= 83; ST11, n= 47; ST656, n= 40) | KPC-2 (n= 184) |
| ***bla*$_{NDM-like}$** | 179 (50.14%) | 14 (ST15, n= 102; ST16, n= 47; ST147, n= 6) | NDM-1 (n= 92)<br>NDM-4 (n= 82)<br>NDM-5 (n=5) |
| ***bla*$_{OXA-like}$** | 82 (22.97%) | 5 (ST16, n=67; ST15, n= 9; ST147, n= 2) | OXA-181(n= 67)<br>OXA-48 (n=15) |
| **Two *bla*-genes** | 165 (46.22%) | 12 (ST15, n= 97; ST16, n= 21, ST147, n=21) | - |
| **Three *bla*-genes** | 55 (15.41%) | 7 (ST16, n= 37; ST15, n=8; ST656, n=2) | - |
| **Four *bla*-genes** | 5 (1.40%) | 1 (ST15, n= 5) | - |

**Supplementary Table 3: The comparison of six circularised plasmid-like sequences carrying *bla*$_{KPC-2}$ gene against the pEC_224 IncN plasmid (NCBI, accession number CP018945).**

| | **27261_8#163** | **27261_8#163** | **26994_7#155** | **26975_5#10** | **27261_8#40** | **27261_8#42** |
|---|---|---|---|---|---|---|
| **Coverage** | 92% | 92% | 98% | 98% | 100% | 100% |
| **Identity** | 99.98% | 99.99% | 99.86% | 100% | 100% | 99.97% |
| **ST15-type** | ST15-17 | ST15-17 | ST15-4 | ST15-19 | NA | ST15-6 |
